# Socioeconomic Status and COVID-19 Related Outcomes in India: Hospital Based Study

**DOI:** 10.1101/2021.05.17.21257364

**Authors:** Arvind Kumar Sharma, Rajeev Gupta, Vaseem Naheed Baig, Teja Veer Singh, Surabhi Chakraborty, Jagdish P Sunda, Prahalad Dhakar, Shiv P Sharma, Raja Babu Panwar, Vishwa Mohan Katoch

## Abstract

**Background & Objective:** COVID-19 infection has disproportionately affected ethnic minorities and deprived populations in Europe and North America. Influence of socioeconomic status on COVID-19 related outcomes has not been studied in India. To determine association of educational status, as marker of socioeconomic status, with COVID-19 related outcomes we performed a study.

**Methods:** Clinically and virologically confirmed successive patients of COVID-19 presenting at a government hospital in India were recruited. Demographic and clinical details were recorded. The cohort was classified according to educational status into Group 1-illiterate or < primary, Group 2-higher secondary, and Group 3-some college. To compare outcomes among groups we performed univariate and multivariate logistic regression and odds ratios (OR) and 95% confidence intervals (CI) were calculated.

**Results:** From March-September 2020 we recruited 4645 patients (men 3386, women 1259) with laboratory confirmed COVID-19. Mean age was 46+18y, most lived in moderate or large households and 30.5% had low educational status. Smoking or tobacco use was in 29.5%, co-morbidities in 28.6% and low oxygen concentration (SpO_2_ <95%) at admission in 30%. Average length of hospital stay was 6.8+3.7 days, supplemental oxygen was provided in 18.4%, high flow oxygen or non-invasive ventilation 7.1%, and mechanical ventilation in 3.6%. 340 patients (7.3%) died. Group 1 patients were younger, more women, larger households, higher tobacco use and were more hypoxic at admission with lower lymphocyte counts, elevated liver enzymes and greater kidney dysfunction. In Group 1 vs Groups 2 and 3 requirement of oxygen (21.6 vs 16.7 and 17.0%), non-invasive ventilation (8.0 vs 5.9 and 7.1%), invasive ventilation (4.6 vs 3.5 and 3.1%) and deaths (10.0 vs 6.8 and 5.5%) were significantly greater (p<0.05). Compared to Group 3, OR for deaths were significantly higher in Group 1 (1.91, 1.46-2.51) and Group 2 (1.24, 0.93-1.66). Adjustment for age, sex, household size, risk factors and comorbidities led to attenuation in OR in Groups 1 (1.44, 1.07-1.93) and 2 (1.38, 1.02-1.85) that remained with adjustments for clinical and laboratory parameters and oxygen support in Groups 1 (1.38, 0.99-1.93) and 2 (1.52, 1.01-2.11).

**Conclusion:** Illiterate and less educational (socioeconomic) status patients with COVID-19 in India have significantly greater adverse in-hospital outcomes and mortality. This is related to more severe disease at presentation.

## INTRODUCTION

COVID-19 pandemic continues to devastate human lives and livelihoods, especially in low and lower-middle income countries.^1^ After the initial spread to the high-income countries in Europe and North America, the epidemic is now rapidly escalating in lower middle and low-income countries of South America, South Asia, South East Asia and Africa.^2^ Epidemiological studies from China, Europe, UK and USA have shown greater disease burden in socioeconomically deprived neighborhoods and minority ethnic groups.^3^ A review that included more than 18.7 million patients from 50 studies in UK and USA reported that individuals from Black and Asian ethnicities had 1.5-2.0 time greater risk of COVID-19 infection compared to White individuals and individuals of Asian ethnicity were at greater risk for intensive-care unit admission and death.^4^ Multiple reasons have been postulated for these socioeconomic disparities and include factors such as poverty, racism and other structural factors, lower availability, access, affordability and utilization of healthcare and low value care.^5,6^ Greater load of infection and longer exposure to the virus due to crowded environments, limited housing, large household sizes, low quality jobs, unsafe commute and undernutrition are also important.^6,7^

Educational status is an important marker of socioeconomic status and hundreds of studies in fields of communicable and non-communicable diseases have reported association of low educational status with adverse health-related events.^8,9,10^ It is also an independent risk factor for morbidity and mortality from infectious diseases.^8,11^ Association of socioeconomic status with COVID-19 related outcomes has not been well studied. A rapid review identified 42 studies that evaluated social determinants of COVID-19 incidence, clinical presentation, health service use and outcomes,^3^ and reported significant associations of race, ethnicity and social deprivation with increased COVID-19 incidence and hospitalization. The review also reported that there was limited evidence regarding other key determinants including occupation, education, housing status and food security and suggested larger epidemiological studies to obtain high-quality evidence. A number of more recent studies have highlighted importance of socioeconomic inequalities in COVID-19 related morbidity and mortality,^12,13,14^ and a review that included 34 studies has reported substantial racial, ethnic and socioeconomic variation in incidence of COVID-19 in USA with greater incidence among poorer communities.^15^

India has one of the largest burdens of COVID-19 cases and deaths.^16^ A macrolevel study reported that Indian states with greater human development index and other socioeconomic indices had higher per capita COVID-19 incidence and deaths.^17^ Although anecdotal evidence and modelling data exist,^1,18^ there are no significant data on association of individual level socioeconomic status with disease incidence and outcomes. Therefore, to examine association of educational status, as a marker of socioeconomic status, in confirmed COVID-19 cases successively admitted to a dedicated COVID-19 government hospital in India we performed a prospective registry-based study.

## METHODS

We conducted a hospital based prospective observational study on patients with laboratory confirmed COVID-19 admitted to a 1200-bed dedicated COVID-19 government university hospital from April to mid-September 2020. Initial data on patients have been reported earlier.^19,20^ The registry has been approved by the college administration and institutional ethics committee (CDSCO Registration Number: CR/762/Inst/RJ/2015). Individual patient consent was waivered by the ethics committee as anonymized data have been used with no patient identifiers. It is registered with Clinical Trials Registry of India at www.ctri.nic.in with registration number REF/2020/06/ 034036.

### Patient data

Successive patients presenting to the hospital for admission with suspicion of COVID-19 infection were enrolled in the study and those who tested positive for COVID-19 on nasopharyngeal and oropharyngeal RT-PCR test were included. A questionnaire was developed and details of sociodemographic, clinical, laboratory, treatments and outcomes variables were recorded using patients’ history and medical files.^19^ Educational status was self-reported and patients were classified into three groups: Group 1-illiterate or < primary education, Group 2-> primary to higher-secondary school education, and Group 3-any graduate or post graduate college education.

### Statistical analyses

The data were computerized and data processing was performed using commercially available statistical software, SPSS v.20.0. Numerical data are expressed as numbers +1 SD and categorical data as percent. Significance of intergroup differences were calculated using either unpaired t-test or χ^2^ test as appropriate. To evaluate association of educational status with clinical outcomes we performed a stepwise logistic regression analysis. Univariate and multivariate odds ratios (OR) and 95% confidence intervals (95% CI) were calculated. P value of <0.05 is considered significant.

## RESULTS

Data were obtained from March 2020 to mid-September 2020 and we enrolled successive patients presenting to the hospital. A total of 7349 patients were hospitalized with confirmed or suspected COVID-19 during this period, 5103 patients (69.0%) tested positive for the disease on reverse transcriptase-polymerase chain reaction (RT-PCR) test and for the present study 4645 individuals (91.0% of confirmed cases), men 3386 (72.9%) and women 1259 (27.1%), in whom detailed clinical data were available have been included (Table 1). The mean age of the cohort was 45.9+18 years, 54% were less than 50 years and about half lived in large family households. Prevalence of low educational status was high and greater in women while tobacco use was more in men (Table 1). Comorbidities were present in 28.6% with hypertension and diabetes being the most common. Details of symptoms, laboratory investigations and clinical status at admission is shown in Table 1. Data on hematological investigations were available in 4456 (95.9%) and for biochemical tests in 867 (18.7%) patients. All patients received standard treatment according to guidelines available from Indian Council of Medical Research and local government.^21^ Management included oral or intravenous hydration, paracetamol and oral or intravenous antibiotics if required. A number of patients also received hydroxychloroquine, ivermectin, azithromycin, doxycycline, lopinavir-ritonavir, favipiravir, etc. The average length of stay in hospital was 6.8+3.7 days, and was significantly greater in men (6.9+3.8 days) than in women (6.5+3.6 days) (p= 0.004). Oxygen requirement was significantly greater in women but other outcomes such as requirement of high flow oxygen, non-invasive or invasive ventilation were not significantly different. Number of in-hospital deaths were significantly greater in men (n=282, 8.3%) as compared to women (n=58, 4.6%) (p<0.001).

**Table 1:** Demographic and Clinical Characteristics of the Study Cohort.

| **Variables** | **Total**<br>**[N=4645]** | **Men (n=3386)** | **Women**<br>**(N=1259)** | **P**<br>**value** |
| --- | --- | --- | --- | --- |
| **Age (mean, yr)** | 45.9±18.0 | 45.5±17.8 | 47.1±18.5 | 0.226 |
| **Age groups** |  |  |  | 0.008 |
| <30 | 1125[24.2] | 838[24.7] | 287[22.8] |  |
| 30-49 | 1397[30.1] | 1052[31.1] | 345[27.4] |  |
| 50-69 | 1650[35.5] | 1161[34.3] | 489[38.8] |  |
| 70+ | 473[10.2] | 335[9.9] | 138[11.0] |  |
| **Family members/house** |  |  |  | 0.834 |
| 1-4 | 2395[51.2] | 656[51.7] | 1739[51.1] |  |
| 5-9 | 2000[42.8] | 535[42.1] | 1465[43.0] |  |
| ≥10 | 281[6.0] | 79[6.2] | 202[5.9] |  |
| **Educational status \*** |  |  |  | 0.000 |
| Illiterate or Primary | 1424[30.5] | 980[28.8] | 444[35.0] |  |
| Secondary school/ Higher secondary | 1538[32.9] | 1061[31.2] | 477[35.0] |  |
| Graduate | 1667[35.7] | 1339[39.3] | 328[25.8] |  |
| **Tobacco or smoking (ever)** | 1369[29.5] | 1045[30.9] | 324[25.7] | 0.001 |
| **Medical co-morbidities** | 1335[28.6] | 1020[29.9] | 315[24.8] | 0.001 |
| Hypertension | 831[17.8] | 658[19.3] | 173[13.6] | 0.000 |
| Pulmonary disease | 193[4.1] | 135[4.0] | 58[4.6] | 0.364 |
| Type 2 Diabetes | 777[16.6] | 666[19.6] | 111[8.7] | 0.000 |
| Thyroid disease | 38[0.8] | 27[0.8] | 11[0.9] | 0.855 |
| Heart disease | 75[1.6] | 51[1.5] | 24[1.9] | 0.360 |
| Neurological disease | 15[0.3] | 6[0.2] | 9[0.7] | 0.008 |
| Current or past tuberculosis | 106[2.3] | 78[2.3] | 28[2.3] | 0.874 |
| Other | 112[2.4] | 55[1.6] | 57[4.5] | 0.000 |
| **Clinical findings** |  |  |  |  |
| Pulse rate /min | 83.9±11.4 | 83.91±11.2 | 84.1±11.8 | 0.715 |
| Systolic BP mmHg | 125.4±12.2 | 125.12±11.9 | 126.0±12.9 | 0.028 |
| Diastolic BP mmHg | 82.8±8.1 | 82.71±7.9 | 83.1±8.4 | 0.155 |
| Respiratory rate/min | 19.0±3.7 | 19.0±3.7 | 19.1±3.9 | 0.313 |
| **SpO<sub>2</sub> at admission** |  |  |  | 0.601 |
| ≥95% | 2144[70.0] | 1554[70.5] | 590[68.7] |  |
| 90-94% | 561[18.3] | 397[18.0] | 164[19.1] |  |
| <90% | 357[11.7] | 252[11.4] | 105[12.2] |  |
| **Laboratory Investigations (Biochemistry n=867; Hematology n=4456)** |  |  |  |  |
| Creatinine, mg/dl | 0.95±0.50 | 0.94±0.47 | 0.97±0.56 | 0.378 |
| SGOT, IU | 44.9±96.5 | 45.0±108.9 | 44.8±44.5 | 0.531 |
| SGPT, IU | 43.4±56.2 | 42.7±59.7 | 45.6± 44.2 | 0.096 |
| Sodium, mEq/L | 136.1±12.5 | 136.6±9.4 | 134.8±17.8 | 0.144 |
| Potassium, mEq/L | 5.4±10.1 | 5.1±8.9 | 5.9±12.6 | 0.112 |
| Hb (gm%) | 12.7±2.3 | 12.7±2.3 | 12.6±2.2 | 0.411 |
| White cells (10<sup>9</sup> cells/L) | 7527±3830 | 7585±3894 | 7370±3651 | 0.099 |
| Lymphocytes (10<sup>9</sup> cells/L) | 1589±1325 | 1607±1355 | 1534±1225 | 0.089 |
| Granulocytes (10<sup>9</sup> cells/L) | 5136±2741 | 5102±2700 | 5241±2864 | 0.118 |
| Lymphocyte/Neutrophil ratio | 0.36±0.32 | 0.36±0.27 | 0.35±0.46 | 0.346 |
| **Outcome measures** |  |  |  |  |
| Mean duration of hospital stay[days] | 6.8±3.7 | 6.9±3.8 | 6.5±3.6 | 0.004 |
| Oxygen requirement | 861[18.4] | 600[17.6] | 261[20.6] | 0.022 |
| High flow O<sub>2</sub>/non-invasive ventilation | 334[7.1] | 236[6.9] | 98[7.7] | 0.371 |
| Mechanical ventilation | 169[3.6] | 123[3.6] | 46[3.6] | 1.000 |
| Recovered | 4217[90.2] | 3020[88.7] | 1197[94.3] | 0.000 |
| Referred | 119[2.5] | 104[3.0] | 15[1.2] | 0.000 |
| Deaths | 340[7.3] | 282[8.3] | 58[4.6] | 0.000 |
| Numbers ± indicate 1 SD; Numbers in parentheses are percent; BP blood pressure; SpO<sub>2</sub> saturation of peripheral oxygen; SGOT serum glutamic oxalate transferase; SGPT serum glutamic pyruvate transferase | | | | |

The cohort was divided into the three groups based on educational status. Important demographic and clinical characteristics and in-hospital outcomes are shown in Table 2. Low educational status (Group 1 and 2) was more common in women while more men had college education. Family size was larger among the less literate group and tobacco use and smoking greater. Prevalence of co-morbidities, especially hypertension and diabetes, was significantly greater among the more literate, similar to previous studies in India.^22^ No significant differences were observed in complaints or clinical findings (data not shown). Low SpO_2_ (<90% as well as <95%), lymphopenia, higher transaminases and higher creatinine values at admission were observed among the less literate. The length of hospital stay was not significantly different in the three groups. Various clinical outcomes are shown in Figure 1 and compared to Group 3, in Group 1 there was greater oxygen requirement (unadjusted OR 1.34, 95% CI 1.12-1.61), non-invasive ventilation (1.14, 0.87-1.49) and invasive ventilation (1.54, 1.06-2.23) (Table 2). Compared to Group 3 (deaths n=92, 5.5%), deaths were significantly greater in Group 1 (n=143, 10.0%, unadjusted OR 1.91, CI 1.46-1,51) and Group 2 (n=104, 6.8%, unadjusted OR 1.24, CI 0.93-1.66) (p<0.001).

**Table 2:**
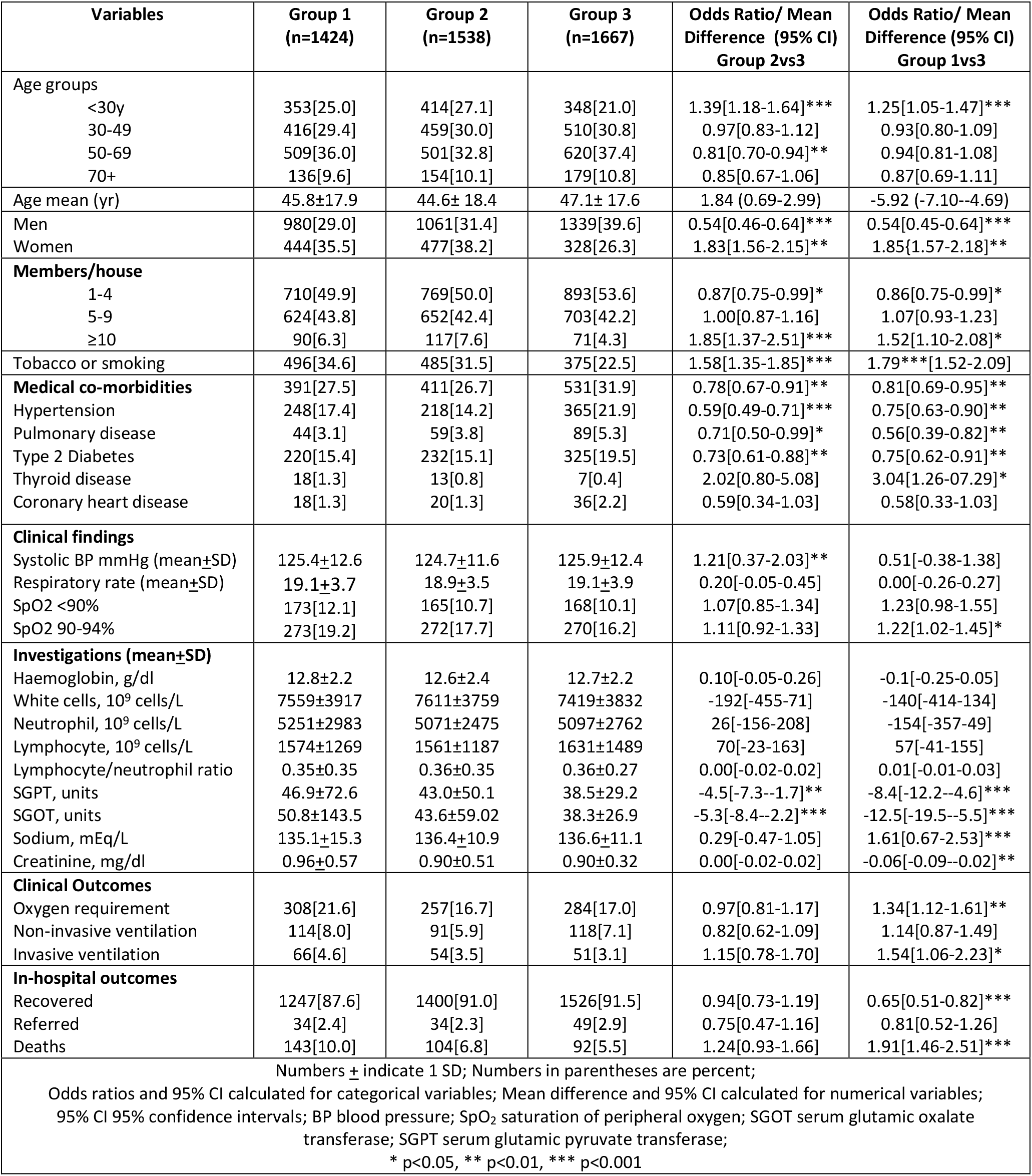
Clinical characteristics and outcomes according to educational status (Group 1= none/primary; Group 2= higher secondary; Group 3= college)

**Figure 1:**
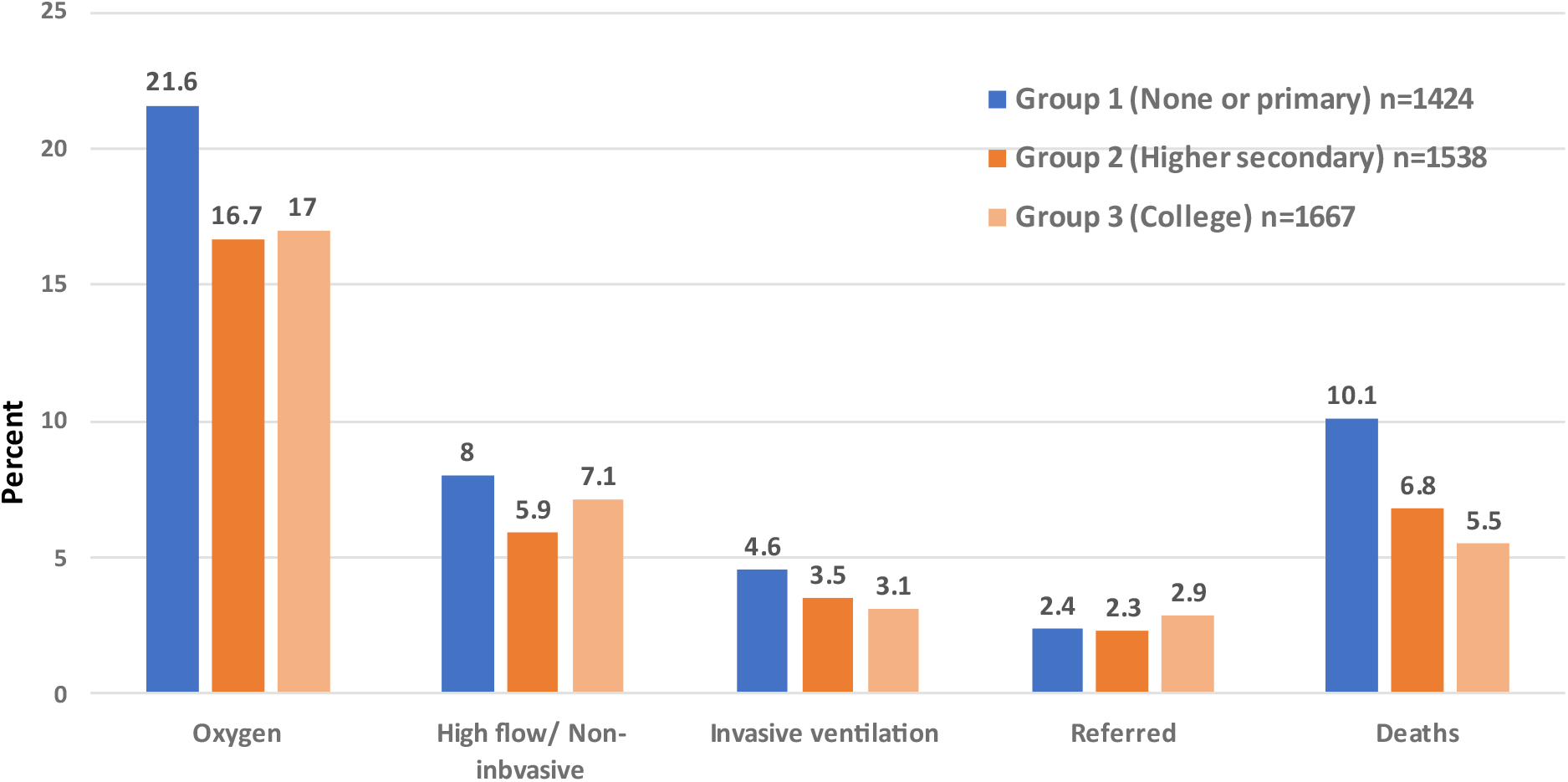
Clinical outcomes in various educational status groups

We performed a stepwise logistic regression analysis to identify influence of various sociodemographic, risk factor, clinical and treatment variables on outcomes. Compared to Group 3, unadjusted OR for deaths were higher in less literate Groups 1 and 2 (Table 3). Following adjustments for age, sex, household size, risk factors and comorbidities the ORs declined but remained significant in both Groups 1 (1.44, 1.07-1.93) and 2 (1.38, 1.02-1.85). However, after addition of clinical features at admission and laboratory investigations the risks attenuated to marginally significant in Group 1 (1.39, 0.99-1.93) and significant in Group 2 (1.53, 1.10-2.11) and remain the same after further adjustments for oxygenation (Table 3). OR for other outcomes assessed in the cohort (need for invasive ventilation and non-invasive ventilation) are shown in Table 3 and demonstrate a marginal significance.

**Table 3:**
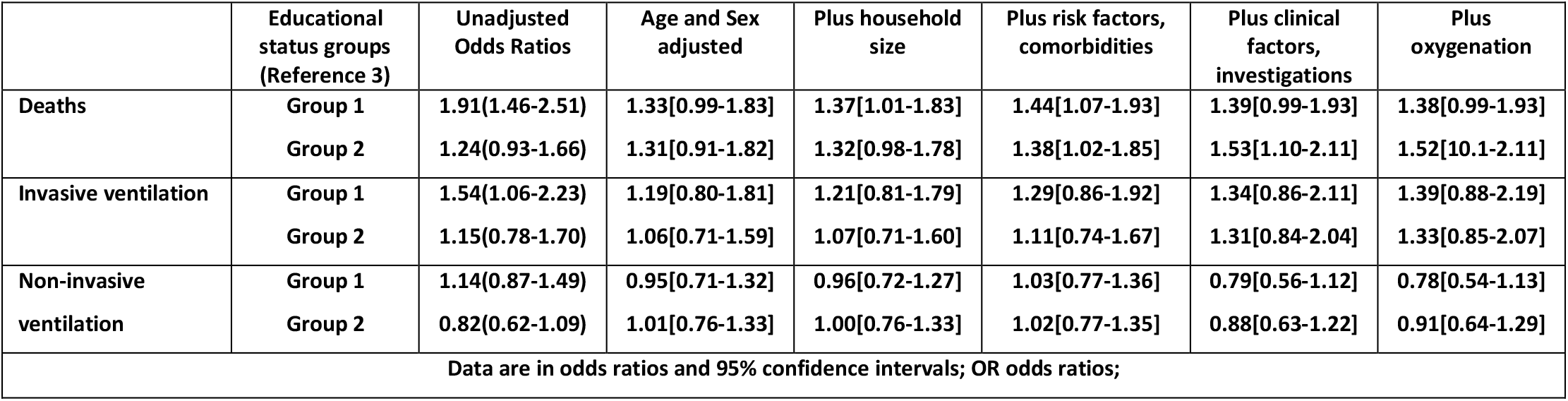
Stepwise Multivariate Logistic Regression Analyses and Odds Ratio for Adverse Outcomes in Educational Status Groups 1 (<primary education) and 2 (>primary-higher secondary education) compared to Group 3 (any college education)

|  | Educational status groups (Reference 3) | Unadjusted Odds Ratios | Age and Sex adjusted | Plus household size | Plus risk factors, comorbidities | Plus clinical factors, investigations | Plus oxygenation |
| --- | --- | --- | --- | --- | --- | --- | --- |
| Deaths | Group 1 | 1.91(1.46-2.51) | 1.33[0.99-1.83] | 1.37[1.01-1.83] | 1.44[1.07-1.93] | 1.39[0.99-1.93] | 1.38[0.99-1.93] |
|  | Group 2 | 1.24(0.93-1.66) | 1.31[0.91-1.82] | 1.32[0.98-1.78] | 1.38[1.02-1.85] | 1.53[1.10-2.11] | 1.52[1.01-2.11] |
| Invasive ventilation | Group 1 | 1.54(1.06-2.23) | 1.19[0.80-1.81] | 1.21[0.81-1.79] | 1.29[0.86-1.92] | 1.34[0.86-2.11] | 1.39[0.88-2.19] |
|  | Group 2 | 1.15(0.78-1.70) | 1.06[0.71-1.59] | 1.07[0.71-1.60] | 1.11[0.74-1.67] | 1.31[0.84-2.04] | 1.33[0.85-2.07] |
| Non-invasive ventilation | Group 1 | 1.14(0.87-1.49) | 0.95[0.71-1.32] | 0.96[0.72-1.27] | 1.03[0.77-1.36] | 0.79[0.56-1.12] | 0.78[0.54-1.13] |
|  | Group 2 | 0.82(0.62-1.09) | 1.01[0.76-1.33] | 1.00[0.76-1.33] | 1.02[0.77-1.35] | 0.88[0.63-1.22] | 0.91[0.64-1.29] |
| Data are in odds ratios and 95% confidence intervals; OR odds ratios; |  |  |  |  |  |  |  |

## DISCUSSION

The study shows that illiterate and less literate COVID-19 patients in India have significantly greater mortality compared to the better educated. The higher risk of death among the less literate persists after adjustment for various sociodemographic factors (age, sex, household size), lifestyle factors and comorbidities but attenuates after adjustment for clinical features at presentation, investigations and oxygen treatment. This suggests that more adverse features at presentation (hypoxia, deranged liver and kidney functions) could be responsible for higher deaths among the less educated (low socioeconomic status) COVID-19 patients in India.

Clinical and epidemiological studies from most developed countries in Europe and North America have consistently reported higher communicable disease-related mortality among the less literate and lower socioeconomic individuals.^11^ In the COVID-19 pandemic, studies from most developed countries have reported greater COVID-19 related mortality and adverse outcomes among the ethnic minorities.^3,4,5^ However, association of mortality among low socioeconomic or less educational status individuals are inconclusive.^3,4,12,13,14^ In England, OpenSAFELY platform evaluated ethnic differences in COVID-19 related hospitalization, intensive care unit admission and death in 17 million adults from the National Health Service.^23^ As compared to British White group, deaths were higher in South Asians in the first wave (OR 1.08, CI 1.07-1.09), and the second wave of COVID-19 epidemic (OR 1.87, CI 1.68-2.07) as well as in the overall cohort (OR 1.26, CI 1.15-1.37). Deaths were the highest in the most deprived groups.^23^ A study from Brazil reported that those with low education attainment were more likely to die from COVID-19 (OR 1.13, CI 1.07-1.19).^24^ Increased deaths among the poor and low educational status patients has also been reported in recent studies from USA,^25^ South Korea,^26^ and African countries.^27^ An epidemiological study in Santiago, Chile report a strong association between socioeconomic status and mortality, measured either by COVID-19 attributed deaths or excess deaths with greater caser-fatality rates in the young people in deprived localities.^28^ Our study is one of the first reports from India that has evaluated socioeconomic difference in COVID-19 related mortality and shows a 1.4 to 1.9 fold greater mortality among low educational status men and women and is similar to the recent international studies. Our study also shows that greater mortality among low educational status individuals could be due to delayed presentation and more severe disease (lower oxygen, greater impaired liver and renal functions) and greater need of oxygen and non-invasive and invasive ventilation in these patients (Table 2). We did not obtain exact information regarding use of various non-evidence based empirical therapies (hydroxychloroquine, ivermectin, lopinavir-ritonavir, favipiravir, etc)^29^ or proven evidence-based therapies such as corticosteroids, remdesivir and tocilizumab,^30^ and this is a study limitation.

A variety of approaches to conceptualization and measurement of socioeconomic status have been used. Four measures are consistently associated with greater risk: low education, low income, lower employment status, and neighborhood socioeconomic factors.^31^ Low education or socioeconomic status is well known as a leading modifiable risk factor for overall as well as infectious disease mortality and is an important social determinant of health.^32^ Previous studies in India and other low and lower middle income countries have reported strong correlation of educational status with measures of income, household wealth, occupation, etc.^33,34^ There are multiple social, clinical and system level contributors that lead to greater disease risk among the poor and include structural barriers to good health, particularly among the less literate and poor, increased risk of exposure (unhygienic working conditions and crowded housing), unequal access to testing and high-quality care, higher rates of associated medical conditions and less access to vaccination.^7^ In the present study we observed some of these barriers among our patients (crowded housing, greater tobacco use, and delayed presentation with more severe disease). COVID-19 in India could act as a catalyst to improve overall healthcare systems with opportunities for policymakers, advocacy groups and researchers for evaluation of various interventions.^36^ It is hoped that COVID-19 would lead to global focus on creation of health equity by influencing coaxing politicians towards the right direction.^37^

The study has some strengths and many limitations. This is the largest case-series from India, we used data from a government hospital which is more representative of general population, there are substantial number of less literate patients reflecting local educational status. This has led to data granularity and robust evaluation of outcomes. Limitations include lack of many sociodemographic factors (occupation, income, working conditions, etc.), clinical parameters (pulmonary findings, radiological evaluation, chest computerized tomographic scans, and blood biomarkers-C-reactive protein, interleukins, d-Dimer, ferritin, etc), and type of therapy the patients received. We also did not evaluate cardiovascular biomarkers (troponins, n-terminal pro-brain natriuretic peptide) that are important in prognostication. There could be multiple causes of deaths in COVID-19 (acute respiratory distress syndrome, myocardial infarction, acute heart failure, pulmonary embolism, sepsis, acute renal failure, etc) and we did not have data on specific causes of death. About 2.5% persons were transferred from our hospital to other centres and although we have obtained information on death in these patients using telephonic interview with families, details of specific outcomes are not available. And finally, data from a single hospital with about 4500 patients and 340 deaths may not be applicable to the whole country which has the second largest burden of COVID-19 in the world.^16^ In view of the massive second wave of COVID-19 in India,^16^ we should strive for larger multicentric studies for identifying reasons for greater mortality among the low socioeconomic status patients with this disease in the country.

In conclusion, our study shows a significantly greater mortality from COVID-19 in less educated (lower socioeconomic status) individuals in India. This is in contrast to the general impression that COVID-19 is more among the middle-class urban population in the country.^18^ Less educated COVID-19 patients have more severe disease at presentation to hospital and need greater oxygen and ventilatory support. Strategies to increase early diagnosis and access to care for these patients are important and should include public health measures for early detection of disease and early referral to treatment centres for appropriate therapeutic measures.

## Data Availability

All the data are in the manuscript.

## REFERENCES

1. Cash R, Patel V. Has COVID-19 subverted global health? Lancet. 2020; 395:1687–8.

2. Dawood FS, Ricks P, Njie GJ, et al. Observations of the global epidemiology of COVID-19 from the pre-pandemic period using web-based surveillance: a cross-sectional analysis. Lancet Infect Dis. 2020; 20:1255–62.

3. Upshaw TL, Brown C, Smith R, Perri M, Ziegler C, Pinto AD. Social determinants of COVID-19 incidence and outcomes: a rapid review. PLoS One. 2021; 16:e0248336.

4. Sze S, Pan D, Nevill CR, et al. Ethnicity and clinical outcomes in COVID-19: a systematic review and meta-analysis. EClinical Med. 2020; 29:100630.

5. Treweek S, Forouhi NG, Venkat Narayan KM, Khunti K. COVID-19 and ethnicity: who will research results apply to? Lancet. 2020; 395:1955–7.

6. Egede LE, Walker RJ. Structural racism, social risk factors and COVID-19-a dangerous convergence for black Americans. N Engl J Med. 2020; 383:e77.

7. Lavizzo-Mourey RJ, Besser RE, Williams DR. Understanding and mitigating health inequities-past, current and future directions. N Engl J Med. 2021; 384:1681–4.

8. Leon DA, Walt G. Poverty, Inequality and Health: An International Perspective. Oxford. Oxford University Press. 2001.

9. Marmot M, Wilkinson R. Social Determinants of Health. Oxford, Oxford University Press. 2005.

10. Gupta R, Joseph P, Rosengren A, Yusuf S. Location and level of care, education, availability of medicines and cardiovascular mortality. In: Fuster V, Narula J, Vaishnava P, Leon MB, Callans DJ, Rumsfeld JS. Editors. Hurst’s The Heart. 15^th^ Ed. New York. McGraw Hill. 2021.

11. Bollyky TJ. Plagues and the Paradox of Progress. Cambridge, MA. The MIT Press. 2018.

12. Gao YD, Ding M, Dong X, et al. Risk factors for severe and critically ill COVID-19 patients; a review. Allergy. 2021; 76:428–55.

13. Liao TF, Maio FD. Association of social and economic inequality with coronavirus disease 2019 incidence and mortality across US counties. JAMA Netw Open. 2021; 4:e2034578.

14. Clouston SAP, Natale G, Link BG. Socioeconomic inequalities in the spread of coronavirus-19 in the United States: a examination of the emergence of social inequalities. Soc Sci Med. 2021; 268:113554.

15. Mackey K, Ayers CK, Kondo KK, et al. Racial and ethnic disparities in COVID-19 related infections, hospitalizations and deaths. Ann Intern Med. 2021; 174:362–73.

16. Ritchie H, Ortiz-Ospina E, Beltekian D, et al. India: Coronavirus Pandemic Country Profile. Available at: https://ourworldindata.org/coronavirus/country/india. Accessed May 5, 2021.

17. Gaur K, Khedar RS, Mangal K, Sharma AK, Dhamija RK, Gupta R. Macrolevel association of COVID-19 with non-communicable disease risk factors in India. Diabetes Metab Syndr: Clin Res Rev. 2021; 15:343–50.

18. Das A, Ghosh S, Das K, Basu T, Das M, Dutta I. Modeling the effect of area deprivation on COVID-19 incidences: a study in Chennai megacity, India. Public Health. 2020; 185:266–269.

19. Sharma AK, Ahmed A, Baig VN, et al. Characteristics and outcomes of hospitalized young adults with mild to moderate COVID-19 at a university hospital in India. J Assoc Physicians India. 2020; 68(8):62–65.

20. Sharma S, Sharma AK, Dalela G, et al. Association of SARS CoV-2 cycle threshold (Ct) with clinical outcomes: a hospital-based study. J Assoc Physicians India. 2021; 69(6):86–90.

21. Government of India, Ministry of Health and Family Welfare. Clinical management protocol: COVID-19. Available at: http://www.rajswasthya.nic.in/PDF/COVID%20-19/FOR%20HOSPITALS/27.06.2020.pdf. Accessed 30 April 2021.

22. Gupta R, Gaur K, Ram CVS. Emerging trends in hypertension epidemiology in India. J Human Hypertens. 2019. 33:575–87.

23. Mathur R, Rentsch CT, Morton CE, et al. Ethnic differences in SARS-CoV-2 infection and COVID-19 related hospitalization, intensive care unit admission, and death in 17 million adults in England: an observational cohort study using the OpenSAFELY platform. Lancet. 2021; 397:1711–24.

24. Li SL, Pereira RHM, Prete CA, et al. Higher risk of death from COVID-19 in low-income and non-White populations of Sao Paulo, Brazil. BMJ Glob Health. 2021; 6:e004959.

25. Azar KMJ, Shen Z, Romanelli RJ, et al. Disparities in outcomes among COVID-19 patients In A large health care system In California. Health Aff (Millwood). 2020; 39:1253–62.

26. Oh TK, Choi JW, Song IA. Socioeconomic disparity and the risk of contracting COVID-19 in South Korea: an NHIS-COVID-19 database cohort study. BMC Public Health. 2021; 21:e144.

27. Salyer SJ, Maeda J, Sembuche S, et al. The first and second waves of the COVID-19 pandemic in Africa: a cross sectional study. Lancet. 2021; 397:1265–75.

28. Mena GE, Martinez PP, Mahmud AS, Marquet PA, Buckee CO, Santillana M. Socioeconomic status determines COVID-19 incidence and related mortality in Santiago, Chile. Science 2021; eabg5298.

29. Siemieniuk RA, Bartoszko JJ, Ge L, et al. Drug treatments for COVID-19: living systematic review and network analysis. BMJ 2020; 370:m2980.

30. RECOVERY: Randomized Evaluation of COVID-19 Therapy. News. Oxford. Nuffield Department of Population Health. 2021. Available at: https://www.recoverytrial.net/news. Accessed 6 May 2021.

31. Braveman P, Egerter S, Williams DR. The social determinants of health: coming of age. Annu Rev Public Health. 2011; 32:381–98.

32. Editorial. Education: a neglected social determinant of health. Lancet Public Health. 2020; 5:e361.

33. Gupta R, Gupta VP, Ahluwalia NS, Educational status, coronary heart disease and coronary risk factor prevalence in a rural population of India. BMJ. 1994; 309:1332–6

34. Gupta R, Kaur M, Islam S, et al. Association of household wealth, educational status and social capital with hypertension awareness, treatment and control in South Asia. Am J Hypertens. 2017; 30:373–381.

35. Smedley BD, Syme SL. Promoting Health: Intervention Strategies from Social and Behavioral Research. Washington. National Academy Press. Institute of Medicine. 2000.

36. Gupta R. Health systems in post-COVID-19 era: strengthening primary care and district hospitals. RUHS J Health Sciences. 2020; 5:61–65.

37. Williams DR, Cooper LA. COVID-19 and health equity-a new kind of herd immunity. JAMA. 2020; 323:2478–80.

